# The REgistry of Flow and Perfusion Imaging for Artificial INtelligEnce with PET (REFINE PET): Rationale and Design

**DOI:** 10.1101/2025.07.10.25330435

**Authors:** Giselle Ramirez, Mark Lemley, Aakash Shanbhag, Jacek Kwiecinski, Robert J.H. Miller, Paul B Kavanagh, Joanna X. Liang, Damini Dey, Leandro Slipczuk, Mark I. Travin, Erick Alexanderson, Isabel Carvajal-Juarez, René R.S. Packard, Mouaz Al-Mallah, Andrew J. Einstein, Attila Feher, Wanda Acampa, Stacey Knight, Viet T Le, Steve Mason, Rupa Sanghani, Samuel Wopperer, Panithaya Chareonthaitawee, Ronny R. Buechel, Thomas L. Rosamond, Robert A deKemp, Daniel S. Berman, Marcelo F. Di Carli, Piotr J. Slomka

## Abstract

**Rationale:** The REgistry of Flow and Perfusion Imaging for Artificial INtelligEnce with PET (REFINE PET) was established to aggregate PET and associated computed tomography (CT) images with clinical data from hospitals around the world into one comprehensive research resource.

**Methods:** REFINE PET is a multicenter, international registry that contains both clinical and imaging data. The PET scans were processed using QPET software (Cedars-Sinai Medical Center, Los Angeles, CA), while the CT scans were processed using deep learning (DL) to detect coronary artery calcium (CAC). Patients were followed up for the occurrence of major adverse cardiovascular events (MACE), which include death, myocardial infarction, unstable angina, and late revascularization (>90 days from PET).

**Results:** The REFINE PET registry currently contains data for 35,588 patients from 14 sites, with additional patient data and sites anticipated. Comprehensive clinical data (including demographics, medical history, and stress test results) were integrated with more than 2200 imaging variables across 42 categories. The registry is poised to address a broad range of clinical questions, supported by correlating invasive angiography (within 6 months of MPI) in 5972 patients and a total of 9252 major adverse cardiovascular events during a median follow-up of 4.2 years.

**Conclusion:** The REFINE PET registry leverages the integration of clinical, multimodality imaging, and novel quantitative and AI tools to advance the role of PET/CT MPI in diagnosis and risk stratification.

## INTRODUCTION

Coronary artery disease (CAD) remains a major public health burden in the United States and worldwide, accounting for healthcare expenditures which are projected to exceed $1 trillion by 2035 [1, 2]. In the wake of increasing rates of obesity, diabetes, and chronic kidney disease, the clinical manifestations of CAD have evolved with a growing proportion of symptomatic patients presenting with predominantly non-obstructive coronary lesions on angiography, often associated with diffuse atherosclerosis and coronary microvascular dysfunction [3–6]. Comprehensive assessment of CAD requires advanced quantitative imaging techniques incorporating information beyond relative perfusion assessments [7, 8] which define only epicardial stenosis. Positron emission tomography (PET) myocardial perfusion imaging (MPI) with associated computed tomography (CT) provides this comprehensive assessment by quantification of myocardial blood flow (MBF) and myocardial flow reserve (MFR) as well as relative perfusion abnormalities. While SPECT remains a more prevalent modality, PET has emerged as the fastest growing cardiovascular imaging test for ischemia over the past decade [9]. Despite this progress, PET MPI continues to be underutilized—not only due to higher capital costs and limited access to PET scanners—but also because of its complexity and the extensive on-site technical expertise needed to fully leverage its comprehensive capabilities, such as MBF analysis. To broaden access for less experienced centers, it is necessary to streamline and automate analysis workflows and to integrate multiparametric data into reporting.

Recognizing these needs – and building on our prior work with the REFINE SPECT registry, which included over 45,000 unique studies [10] and enabled the development and validation of novel SPECT MPI analysis methods (resulting in more than 40 collaborative publications to date) – we established a new multi-center imaging registry for cardiac PET: the REgistry of Flow and Perfusion Imaging for Artificial INtelligEnce with PET (REFINE PET). This registry is designed to serve as a resource for developing, testing, and validating new approaches to PET MPI – with the goal of unlocking its full clinical potential and facilitating broader adoption. This manuscript describes the rationale and design of the REFINE PET registry, including the data collection procedures, quality control (QC) measures, and baseline characteristics of the patients collected to date.

## MATERIALS AND METHODS

### Overall Study Design

We developed a multicenter, international registry to collect and analyze retrospective clinical, procedural, imaging, and follow-up data of patients undergoing PET/CT MPI. The registry included contributions from 14 sites across the United States, Mexico, Canada, Switzerland, and Italy, with participating centers routinely performed MBF quantification and CT attenuation correction. Consecutive PET/CT scans from each site were included to reflect real-world clinical practice. This registry will support validation of both standard and novel image analysis methods, which can help streamline and enhance this cardiovascular modality, for broader clinical use.

The registry employed a collaborative model in which participating sites contributed both clinical and imaging datasets, reflecting routine care at each center, as illustrated in **Figure 1**. These data were harmonized and integrated into a comprehensive clinical-imaging database at the central data coordinating center and core laboratory at Cedars-Sinai Medical Center (CSMC). The database included detailed demographic, clinical and laboratory information, along with PET/CT imaging, corresponding coronary angiography and follow-up data, and deep learning derived coronary calcium data for patients who underwent a PET/CT MPI exam. This integrated structure enabled large-scale, automated image analysis using advanced computational methods, including artificial intelligence.

**Figure 1:**
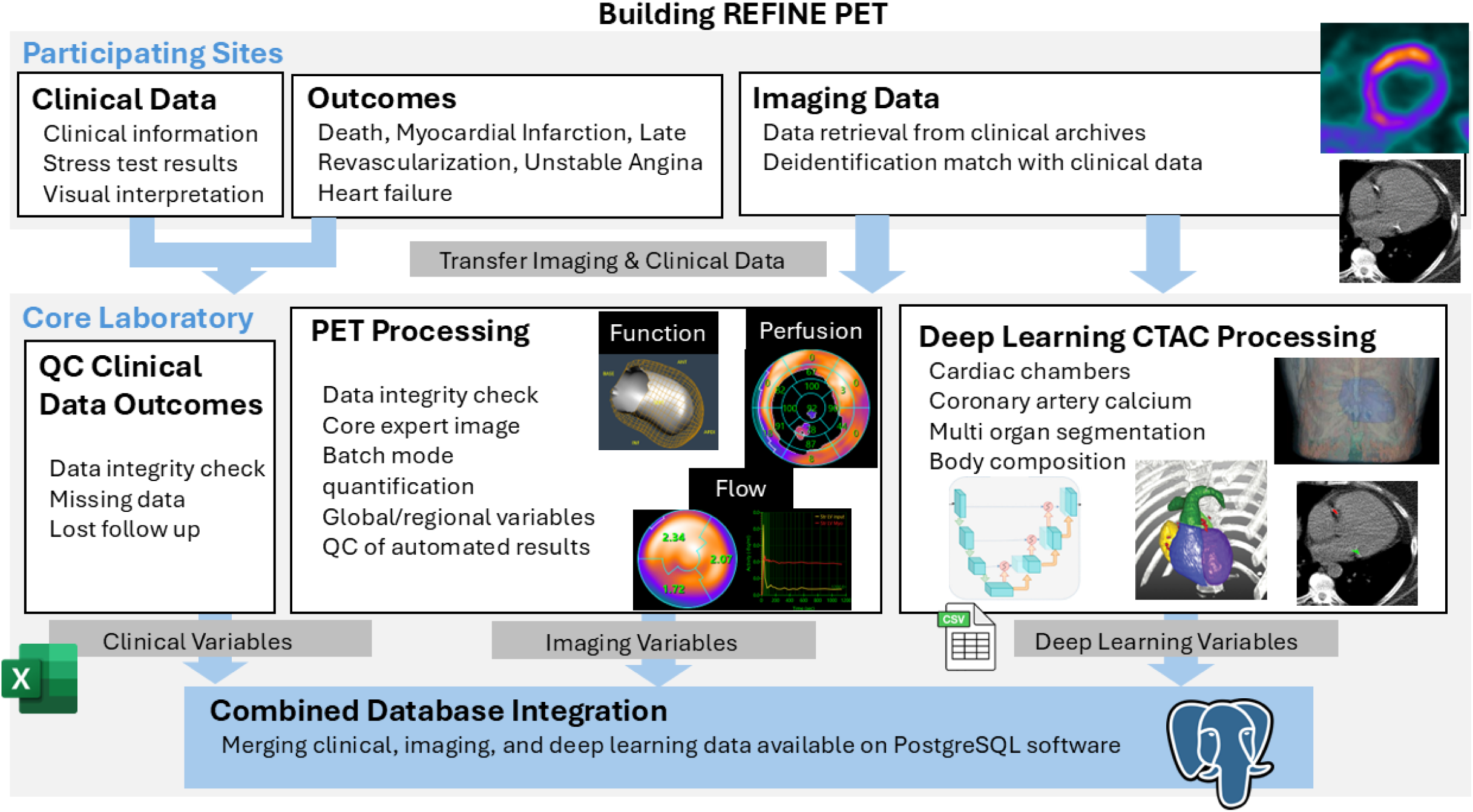
Process of data collection and processing for REFINE PET registry. CTAC: computed tomography attenuation correction; QC: quality control.

This study was approved by the institutional review boards at each participating site. Approval of the overall study was provided by the CSMC institutional review board. The study complied with the Declaration of Helsinki, and sites either obtained written informed consent or a waiver of consent for the use of the deidentified data.

### Study Population

Consecutive patients at each center who underwent PET MPI for evaluation of suspected or known CAD were included. For patients with multiple scans performed on different dates, only the first scan was included in the registry.

The REFINE PET registry currently includes scans performed between 2007 and 2024. Clinical variables collected from each site are summarized in **Table 1**. Dedicated de-identification software, fully compliant with the Health Insurance Portability and Accountability Act (HIPAA), was used to anonymize all data prior to transfer. The core laboratory did not receive any information that could directly or indirectly identify individual patients.

**Table 1:** Clinical variables in REFINE PET:

| Type | Variable |
| --- | --- |
| **General Clinical Variables** | Site |
|  | Patient Location (Inpatient, Outpatient, Emergency) |
|  | Age, Gender |
|  | Weight (kg), Height (cm), BMI (kg/m<sup>2</sup>) |
|  | Clinical Indications for test |
|  | Family History of Coronary Artery Disease |
|  | Anginal Presenting Symptoms |
|  | Race, Ethnicity |
| **Medical History** | Hypertension |
|  | Diabetes Mellitus |
|  | Dyslipidemia |
|  | Currently smoking |
|  | Current medications |
| **History of Heart Disease** | Past MI |
|  | Past PCI/CABG |
|  | Past TAVR |
|  | Past Cardiac Transplant |
|  | Past Other open-heart surgery |
|  | PAD/PVD |
| **Stress variables** | Pharmacological stress agent |
|  | Resting Heart Rate (bpm) |
|  | Peak Stress Heart Rate (bpm) |
|  | Resting BP: Systolic, Diastolic (mmHg) |
|  | Peak Stress BP: Systolic, Diastolic (mmHg) |
|  | Rest ECG |
|  | LVH |
|  | Conduction Disease |
|  | ECG Response to Stress |
|  | Clinical Response to Stress |
|  | ECG ST Deviation (mm), direction, and slope |
| **Clinical interpretation** | Quality of MPI Study |
|  | Visual Perfusion Assessment |
|  | SSS, SRS, SDS |
| **ICA** | % stenosis in coronary arteries |
| **Outcome Variables** | ACM, MI, UA, Revasc, HF, Cardiovascular Death, Follow-up time |
| **Biomarkers** | eGFR |
|  | LDL |
|  | HDL |
|  | cTnT |
|  | cTnI |
|  | BNP |
|  | NT-proBNP |

ACM: all-cause mortality; BMI: body mass index; BNP: B-type natriuretic peptide; BP: blood pressure; CABG: coronary artery bypass grafting; cTnI: cardiac troponin I; cTnT: cardiac troponin T; eGFR: estimated glomerular filtration rate; HDL: high-density lipoprotein; LDL: low-density lipoprotein; LVH: left ventricular hypertrophy; MI: myocardial infarction; NT-proBNP: N-terminal pro-B-type natriuretic peptide; PAD: peripheral arterial disease; PCI: percutaneous coronary intervention; PI: perfusion index; PVD: peripheral vascular disease; SDS: summed difference score; SRS: summed rest score; SSS: summed stress score; TAVR: transcatheter aortic valve replacement; UA: unstable angina; HF: heart failure; ICA: invasive coronary angiography.

### Data Collection

All protected health information (PHI) including date of birth, name, medical record numbers, and imaging dates of imaging scans were removed using image deidentification software. After deidentification, patient scans and corresponding clinical data were securely transferred to the core laboratory at CSMC for quantification and analysis using HIPAA compliant storage systems. Each participating site maintained a cross-referenced master list linking anonymized codes to original identifiers, which was used only to resolve discrepancies. Neither CSMC nor the REFINE PET registry had access to or maintained linkage with these master sheets.

### Diagnostic Outcomes

The diagnostic population comprised patients who underwent invasive coronary angiography (**ICA**) within 180 days of the PET scan. ICA results and revascularization information were collected from medical records using site-specific methods. Stenosis severity per coronary artery segment was determined by an interpreting cardiologist at the time of clinical interpretation of ICA. A stenosis severity of ≥70% in a major coronary artery vessel or its major branches, or ≥50% stenosis severity in the left main coronary artery was considered significant for obstructive CAD.

### Prognostic Outcomes

The prognostic cohort comprised all patients with known or suspected CAD. The primary endpoint for the prognostic cohort was a composite of major adverse cardiac events (MACE), comprising all-cause mortality, non-fatal myocardial infarction (MI) (MI types 1, 4, or 5 according to the fourth universal definition of MI) , admission for unstable angina (UA) (hospitalization for new or worsening cardiac chest pain without evidence of myocardial injury), hospitalization for heart failure (HF) , and coronary revascularization (percutaneous coronary intervention [PCI], or coronary artery bypass grafting [CABG]) with more than 90 days from PET scan. Early revascularization was defined as revascularization within 90 days after the PET scan, while late revascularization was any revascularization occurring post 90 days. Non-fatal MI was defined as worsening chest pain or the recent onset with high cardiac enzyme levels and ischemic electrocardiogram charges that result in hospital admission. Unstable angina was defined as evidence of myocardial ischemia based on diagnostic testing or hospitalization due to recent onset or worsening chest pain [14]. Hospitalization for HF was defined by the primary discharge diagnosis, determined through previously validated International Classification of Diseases (ICD) codes. Cardiovascular deaths included those attributed to myocardial infarction, sudden cardiac death, heart failure, stroke, cardiovascular procedures, cardiovascular hemorrhage, or other cardiovascular causes [15]. Cardiovascular death was determined by review of the death certificate, hospital chart, or physician’s records.

### Site Event Adjudication

Follow-up for MACE and HF was performed locally at each participating center. Non-fatal MI and UA were defined based on hospital admission for recent onset chest pain and admission diagnostics and MI by elevated cardiac enzyme levels and ischemic ECG changes [11, 16]. All-cause mortality determination methods differ by country. In the United States, all-cause mortality was retrieved from the Social Security Death Index and was combined with MACE, while HF was obtained from hospital electronic medical records or patient contact. In Mexico, all-cause mortality was obtained by chart review at the site or from patient contact. In Canada, all-cause mortality was determined by chart review at the institute and physician’s office. In Europe, MACE and HF were obtained from the hospital electronic medical records. ICA results, revascularization details, and data regarding hospitalization due to unstable angina or MI were collected by review of medical records (including all clinics, cardiology groups, and hospital visits). For each patient considered to have a MACE, medical records regarding non-fatal events were reviewed and verified for correctness by site investigators.

### PET Imaging Acquisition and Reconstruction

Patients were scanned in the supine position and images were reconstructed with and without attenuation correction for the PET/CT MPI systems for both rest and stress. The registry included 24 patients who underwent stress-only imaging. Reconstructed images were generated using vendor-recommended iterative reconstruction algorithms optimized for each scanner and site.

### Computed Tomography Imaging Acquisition and Reconstruction

CTAC scans associated with PET scans, including gated coronary artery calcium scans if available, were collected as part of the registry. CT images were acquired with site-specific imaging protocols as outlined in **Supplementary Table 1**.

### Clinical MPI Scoring

At each site, PET/CT scans were evaluated during clinical reporting by reporting clinicians with access to all available data, including electrocardiographic, hemodynamic, stress and rest TPD, gated functional, MBF, MFR, and other clinical data. Clinicians would rate the patient from all available clinical perfusion findings on a five-point scoring system: 0 (normal), 2 (probably normal), 1 (equivocal), 3 (abnormal), and 4 (definitely abnormal) [17]. Apart from West Los Angeles Veterans Affair Medical Center, experienced clinicians visually evaluated the PET perfusion images and performed visual segmental scoring following the 17-segment American Heart Association model [18]. The summed stress score (SSS), summed rest score (SRS), and its difference (summed difference score [SDS]) were collected from sites.

### Imaging Database

All PET and CT images were transferred to the core laboratory, where they were readily accessible for direct and advanced processing. Participating centers transferred anonymized ZCM files (zipped DICOM images) using secure methods such as the Box web server (a cloud service) or other site-approved secure transfer software. This approach ensured the confidentiality and integrity of the data during transmission. The ZCM files, compressed versions of the large PET/CT MPI DICOM data, were roughly half the size of the original files, making storage more efficient. Additionally, ZCM eliminated inherent inefficiency of handling thousands of DICOM images per patient. Upon receipt, the de-identified image data underwent verification for completeness before being loaded into the REFINE PET imaging database (Multidax). Once integrated into Multidax, a quality control process was conducted to ensure the imaging data adhered to established standards, thus maintaining the reliability and accuracy of subsequent analyses.

### Core Laboratory Processes

#### Quality Control of Imaging Data and Data Cleaning of Imaging

Experienced core laboratory technologists, blinded to the clinical data, conducted a thorough review of the PET/CT MPI images. The quality control (QC) process encompassed several important checks, including identifying duplicate imaging files, detecting corrupt imaging files, and locating missing imaging files. Collaboration with each site was essential to reconcile any issues related to corrupt images or missing data, ensuring complete imaging datasets.

The QC process also involved manual correction of automatically generated myocardial segmentation as necessary, visualization of PET/CT MPI registration to ensure proper alignment and accuracy, and the application of fully automated motion correction for dynamic flow using the quantitative PET package QPET (Cedars-Sinai, Los Angeles) and the latest optimized tools. Technologists meticulously documented any processing errors or technical issues encountered with the images. All manual corrections made to the images were automatically recorded by the software and stored within the Multidax image database.

For CT calcium scoring with dedicated gated CT used for coronary artery calcium (CAC) scans, the QPET calcium scoring module was employed to annotate both coronary and extra-coronary calcium.

### Quantification with Clinical Software

After quality control, PET images were quantified in QPET using batch-processing mode. The batch mode quantified all image datasets for all patients sequentially in an automated mode, optimizing the computational resources required to process the image registry. The software (QPET) produced a single row of the image data variables quantified per image dataset. Multiple rows per patient corresponded to static and dynamic gated and CT studies for stress and rest acquisitions. Subsequently, the image data was reshaped by the database to list one unique row per patient, consolidating the patient’s perfusion and functional variables– with any missing variables set to blanks. Combined perfusion flow and functional parameters (computed from stress and rest) were also included.

### CT Processing with Deep Learning

We utilized our previously validated deep learning model for coronary artery calcium (CAC) segmentation on CTAC scans [19]. CAC scoring was then performed according to established methods[20]. In brief, the system consists of two networks, the first of which was trained for segmentation of the heart silhouette and the second network was trained to segment the CAC. A supervised learning regimen was used for both segmentation networks. The heart mask was applied to the final CAC prediction to reduce any spurious bone overcalling or calcification in non-cardiac regions. To imitate the physician approach of aggregating information from adjacent slices, three slices (target slice and two adjacent slices) were provided to both the networks as input. The model includes a correction factor for slice thickness, ensuring consistent scoring despite differences in slice thickness.

### PET Imaging Variables

Imaging variables were calculated at stress and rest for perfusion, gated, and dynamic images, analyzed both on an overall and by per-segment, per-vessel, or per-wall when applicable. In total, 42 imaging variable categories, including general imaging, perfusion, flow, functional, and CT parameters, were automatically quantified **(Table 2)**. These variable categories include separate values for stress/rest, for static, gated, and flow images incorporating the three regional (left anterior descending, left circumflex, right coronary artery) and 17-segment myocardial models. For each patient, this resulted in over 300 variables when using the three vessel regions, and over 2200 variables when including the 17-segments.

**Table 2:** Imaging Variables

| Type | Variable | Stress and Rest | Regional/ Segmental* |
| --- | --- | --- | --- |
| **General Imaging Variables** | **All derived by QPET** |  |  |
|  | Myocardial Counts | ✓ |  |
|  | LV Segmentation QC Metrics | ✓ |  |
|  | LV Dimensions (mm) | ✓ |  |
|  | Myocardial Mass (g) | ✓ |  |
|  | LV Shape Index, Eccentricity | ✓ |  |
|  | Injected Dose (rest and stress, MBq) | ✓ |  |
| **Perfusion Variables** | **All derived transmural/subendocardial by QPET** |  |  |
|  | Total Perfusion Deficit (%) | ✓ |  |
|  | Perfusion Severity | ✓ | ✓ |
|  | Perfusion Defect Extent | ✓ | ✓ |
|  | Stress-Rest Change |  | ✓ |
|  | Segmental Scores | ✓ | ✓ |
|  | Normalized Raw Perfusion Uptake | ✓ | ✓ |
|  | Ejection Fraction (%) | ✓ |  |
| **Functional Variables** | **All derived by QPET** |  |  |
|  | Volumes: EDV, ESV (ml) | ✓ |  |
|  | Ventricle Length: Diastolic, Systolic (mm) | ✓ |  |
|  | Motion (mm) | ✓ | ✓ |
|  | Motion Defect Extent (%) | ✓ | ✓ |
|  | Motion Score | ✓ | ✓ |
|  | Thickening (mm) | ✓ | ✓ |
|  | Thickening Defect Extent (%) | ✓ | ✓ |
|  | Thickening Score | ✓ | ✓ |
|  | Phase SD, Dyssynchrony, Bandwidth, Entropy | ✓ |  |
|  | Diastolic Parameters: PER (EDV/s), PFR (EDV/s), MFR (EDV/s), TTPF (msec) | ✓ |  |
|  | Average RR in ECG/ Beats Per Minute | ✓ |  |
|  | Transient Ischemic Dilation |  |  |
| **Dynamic Variables** | **All derived with/without AMC and RAS transmural/subendocardial by QPET** |  |  |
|  | Myocardial Flow | ✓ | ✓ |
|  | Spill over Fraction | ✓ | ✓ |
|  | Myocardial Flow Reserve | ✓ | ✓ |
|  | CFC/IMFR | ✓ | ✓ |
| **Computed Tomography** | **All derived by deep learning** |  |  |
|  | Agatston Score | ✓ | ✓ |
|  | Volume | ✓ | ✓ |
|  | Density of calcified coronary plaques | ✓ | ✓ |
|  | Number of vessels involved | ✓ | ✓ |
|  | Number of calcified coronary plaques | ✓ | ✓ |

\*Variables were also computed for the 17-segment AHA subdivisions, coronary artery territories (left anterior descending, left circumflex, and right coronary artery), and for myocardial walls (apical, lateral, inferior, septal, and anterior).
AMC: auto motion correction; CFC: coronary flow capacity; EDV: end-diastolic volume; ESV: end-systolic volume; iMFR: integrated myocardial flow reserve; LV: left ventricular; MFR: mean filling rate; PER: peak ejection rate; PFR: peak filling rate; QC: automatic quality control; RAS: residual activity subtraction; RR: rest-to-stress heart rate ratio; SD: standard deviation; TTPF: time to peak filling.

Additionally, both transmural and subendocardial flows were derived to assist in the evaluation of global perfusion defects, subclinical ischemia, microvascular dysfunction, and balanced ischemia.

### Clinical Data Cleaning and Quality Control

The data received from the centers were verified to ensure that all required information had been provided. Additional efforts were made to standardize all possible mismatches between centers regarding the coding of the clinical information. Codes used to report ICA findings, definitions of categorical variables, and encoding of missing data elements were verified. Moreover, the clinical data were checked against quantified image data (one record for every image). Any inconsistencies were resolved with the investigative centers.

### Combined Database

To help the collaboration and ensure the durability of patient records, the clinical data and derived imaging variables were stored in an Object-Relational database system: PostgreSQL. After collecting and cleaning the data from a clinical site, the information was integrated in the PostgreSQL server as a spreadsheet file in a CSV format. This was done periodically when a substantial number of cases were delivered from collaborating sites (typically 1-3 times per site). A dedicated computer ran a PostgreSQL(v12) server in the core laboratory at Cedars-Sinai. The database tables stored the information of each patient in one row. The site name and the anonymized patient code were used as unique identifiers to link clinical data with the image database. Server administrators managed user accounts with passwords to grant access to the database with read and/or write permissions. Any major statistical software (such as R, Stata, SPSS) with an additional ODBC driver package could retrieve, visualize, and analyze information from a PostgreSQL database. An ODBC driver used the Open Database Connectivity interface by Microsoft that allowed applications to access data in the database management systems using SQL (Structured Query Language). Specific views of the database in CSV format could be extracted for analysis purposes.

### Verification of the Combined Clinical-Image Database Integrity

Combined data integrity was subsequently verified at the core lab. For the imaging data, intra-patient studies were verified for mismatches in sex and mismatches in patient age. Patients with mismatches in DICOM’s header and clinical data’s sex and age were followed up with the specific center, resolving any discrepancies.

## Data Confidentiality

The information was coded, such that it did not contain any protected patient information, thus rendering impossible the identification of the patient at the central core laboratory, or by any of the investigators. The core laboratory did not receive or store any information that would directly or indirectly identify the patient corresponding to the data received for the study. All HIPAA requirements were met. DICOM PET MPI and CT images were deidentified by the automated de-identification software, DicoNymizer (Cedars-Sinai, Los Angeles) in batch mode at the collaborating sites before being sent to the core laboratory. No manual de-identification was performed. All project staff members were trained in confidentiality and research ethics principles.

## Statistical Analysis

Continuous variables were presented as medians with interquartile range (IQR; IQ1-IQ3). Categorical variables were summarized using counts and relative frequencies (%). Pearson’s χ2 test was used to compare categorical variables, whereas the Kruskal Wallis test was applied for continuous variables when appropriate. Confidence intervals were estimated using the percentile bootstrap method. A two-tailed p-value of <0.05 was considered statistically significant. Statistical analyses were performed with Pandas (version 2.1.1), Numpy (version 1.24.3), Scipy (version 1.11.4), and Scikit-learn (version 1.3.0) in Python version 3.11.5 (Python Software Foundation, Wilmington, DE, USA), as well as R studio (R version 4.4.1 Inc, Boston, MA, USA).

## RESULTS

### Image Processing

The processing time in batch-mode per patient was around 2 seconds. The total storage requirement to date for the final ZCM files is 9.1 TB.

### Patient Characteristics

The current REFINE PET registry comprises 35,588 patients with suspected or established CAD from 14 sites, including 21,010 (59%) males; a median age was 68 (IQR 59-76) **(Table 3)**. The clinical and imaging quantification database holds combines clinical and imaging data as tabular variables.

**Table 3:** Baseline Characteristics

| **Characteristic** | **Overall**<br>**N = 35,588<sup>1</sup>** | **With ICA results**<br>**N = 5,971<sup>1</sup>** |
| --- | --- | --- |
| Age | 68 (59, 76) | 69 (61, 76) |
| Male | 21,010 (59%) | 4,159 (70%) |
| BMI (kg/m<sup>2</sup>) | 29.1 (25.2, 34.4) | 29.1 (25.5, 34.1) |
| BMI (kg/m<sup>2</sup>) <25 | 8047 (23%) | 2,658 (45%) |
| BMI (kg/m<sup>2</sup>) 25-30 | 11547 (32%) | 5,001 (84%) |
| BMI (kg/m<sup>2</sup>) >30 | 15788 (44%) | 4,728 (80%) |
| Diabetes Mellitus | 12,635 (36%) | 1,048 (18%) |
| Hypertension | 27,538 (78%) | 1,764 (30%) |
| Dyslipidemia | 25,897 (74%) | 1,138 (20%) |
| Smoking | 5,929 (17%) | 3,022 (51%) |
| Family History | 10,036 (29%) | 69.0 (61.0, 76.0) |
| PVD | 4,589 (14%) | 4,159 (70%) |
| Known CAD | 11,580 (33%) | 29.1 (25.5, 34.1) |
| CAC score* | 140.8 (0.0, 984.4) | 633.0 (82.1, 1,961.3) |
| CAC > 0* | 17,861 (71%) | 3,878 (86%) |
| Stress TPD* | 4.1 (1.5, 10.2) | 12.6 (6.0, 22.6) |
| Rest TPD* | 0.6 (0.0, 2.9) | 2.5 (0.5, 7.9) |
| Stress EF* | 67.8 (56.0, 75.7) | 58.7 (44.1, 68.6) |
| Rest EF* | 64.6 (53.6, 72.5) | 58.1 (44.2, 67.9) |
| Stress MBF* | 2.2 (1.6, 2.8) | 1.7 (1.3, 2.2) |
| Rest MBF* | 0.9 (0.7, 1.2) | 0.9 (0.7, 1.1) |
| MFR* | 2.3 (1.8, 2.9) | 2.0 (1.5, 2.5) |
| **Outcomes** |  |  |
| Early Revasc (≤90 days) | 2,795 (7.9%) | 2,517 (42%) |
| Death | 6,235 (18%) | 1,469 (25%) |
| MI | 2,122 (6.0%) | 729 (12%) |
| Late PCI (>90 days) | 1,742 (5.4%) | 661 (12%) |
| Late CABG (>90 days) | 380 (1.1%) | 133 (2.2%) |
| Unstable Angina | 990 (4.4%) | 467 (12%) |
| MACE | 9,252 (26%) | 2,531 (42%) |
| Heart failure admission | 2,439 (8.6%) | 697 (14%) |

<sup>1</sup>Median (Q1, Q3); n (%)
\*Based on patients with cleaned imaging and clinical data and with available CTAC, stress perfusion, and stress dynamic (n=25309) and ICA cohort of that group (n=4484)
CABG: coronary artery bypass grafting; CAC: coronary artery calcium; CAD: coronary artery disease; EF: ejection fraction; ICA: invasive coronary angiography; MACE: major adverse cardiovascular event; MBF: myocardial blood flow; MI: myocardial infarction; PCI: percutaneous coronary intervention; PVD: peripheral vascular disease; TPD: total perfusion deficit.

### Imaging protocols

The imaging protocols for all sites are listed in **Table 4**.

**Table 4:**
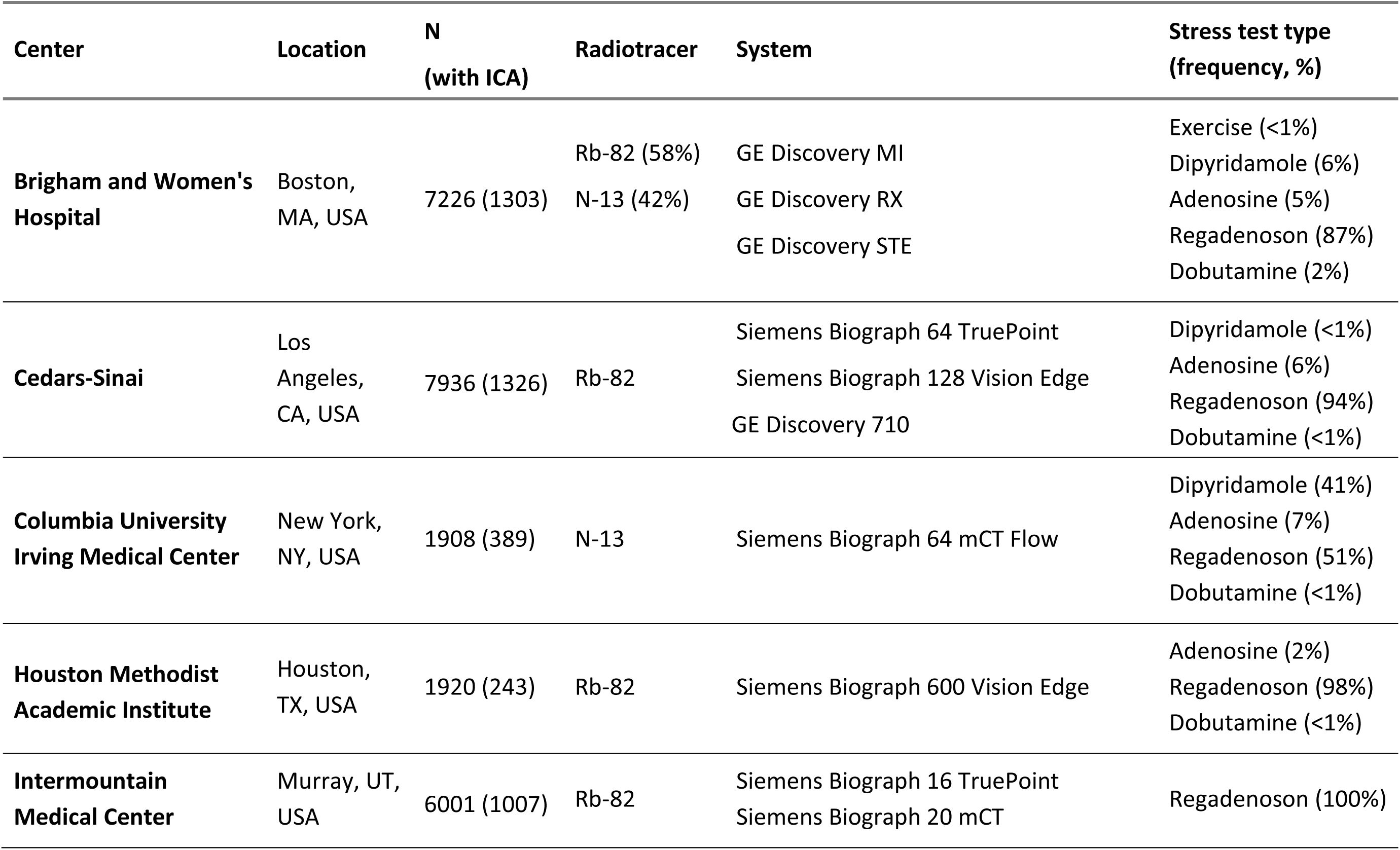
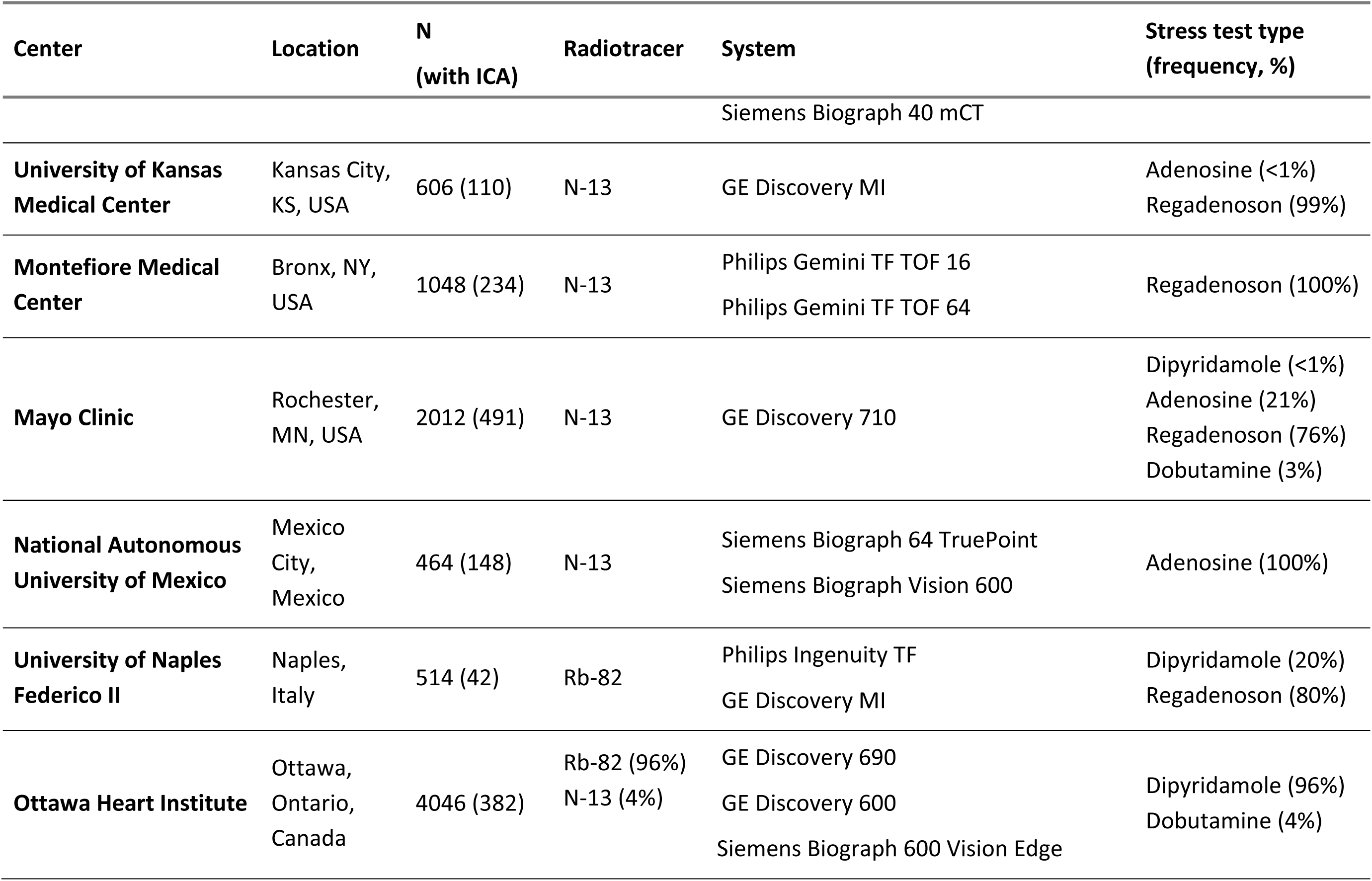
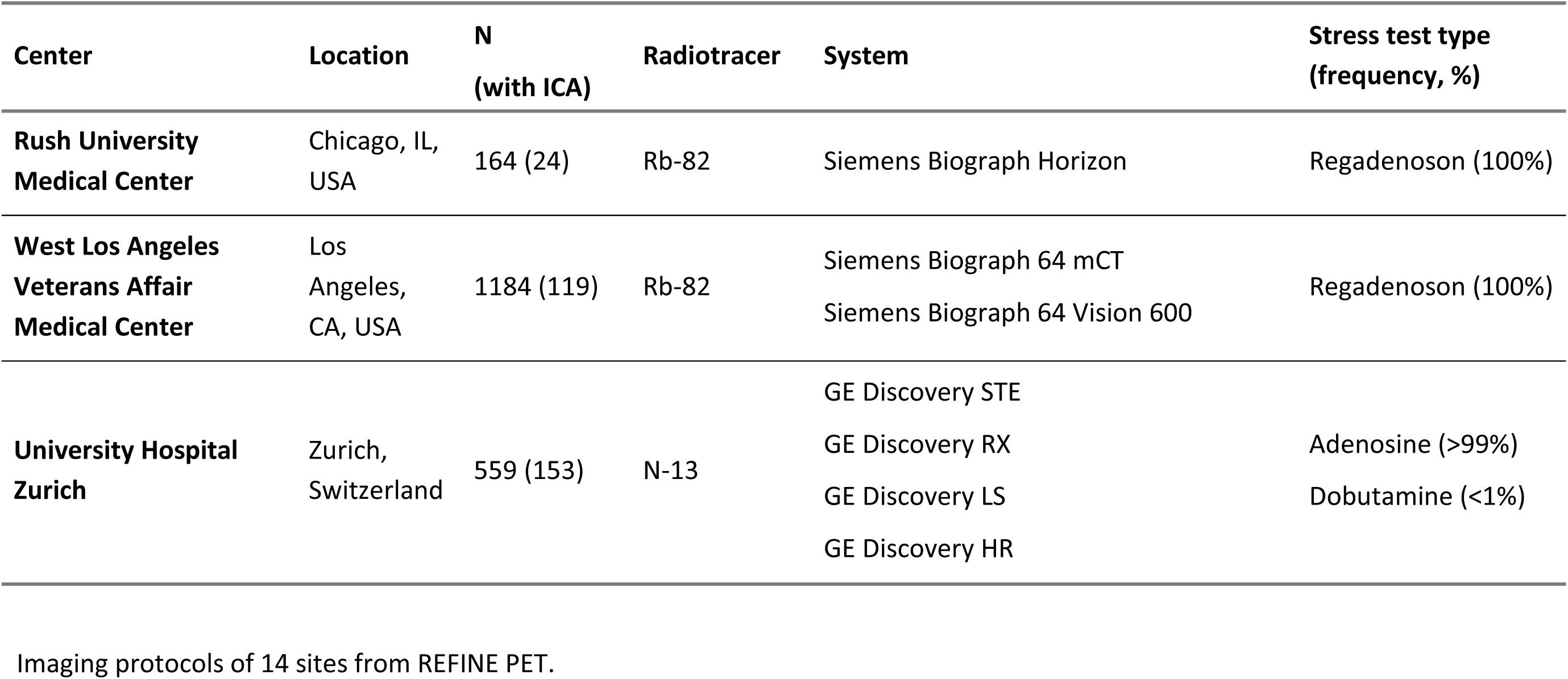
Imaging Protocols

### Diagnostic Outcomes

The registry currently includes 5971 patients (17%) who underwent ICA within 6 months of their PET scan **(Table 3)**. Detailed angiographic data are still being checked for 944 patients. Of the 5027 patients with complete and verified angiographic data, 3009 (60%) had obstructive CAD.

### Prognostic Outcomes

The registry currently has 9252 patients (26%) MACE events during a median follow-up time of 4.2 years (IQR 2.4 – 5.6 years). Among patients who experienced MACE, 2531 underwent ICA within 6 months of the PET scan. These include 6235 (18%) deaths, 2122 (6%) non-fatal myocardial infarctions, 2082 (6%) underwent late revascularizations, and 990 (3%) admissions for unstable angina. There were also 2439 (7%) admissions for HF. A comparison between the overall population and patients who underwent ICA within 6 months of PET is shown in **Table 3**.

An overview of the prognostic outcomes and its relation to calcium, perfusion, and MFR variables for the data that has undergone QC, along with all available clinical data received from the site, is shown in **Figure 2**.

**Figure 2:**
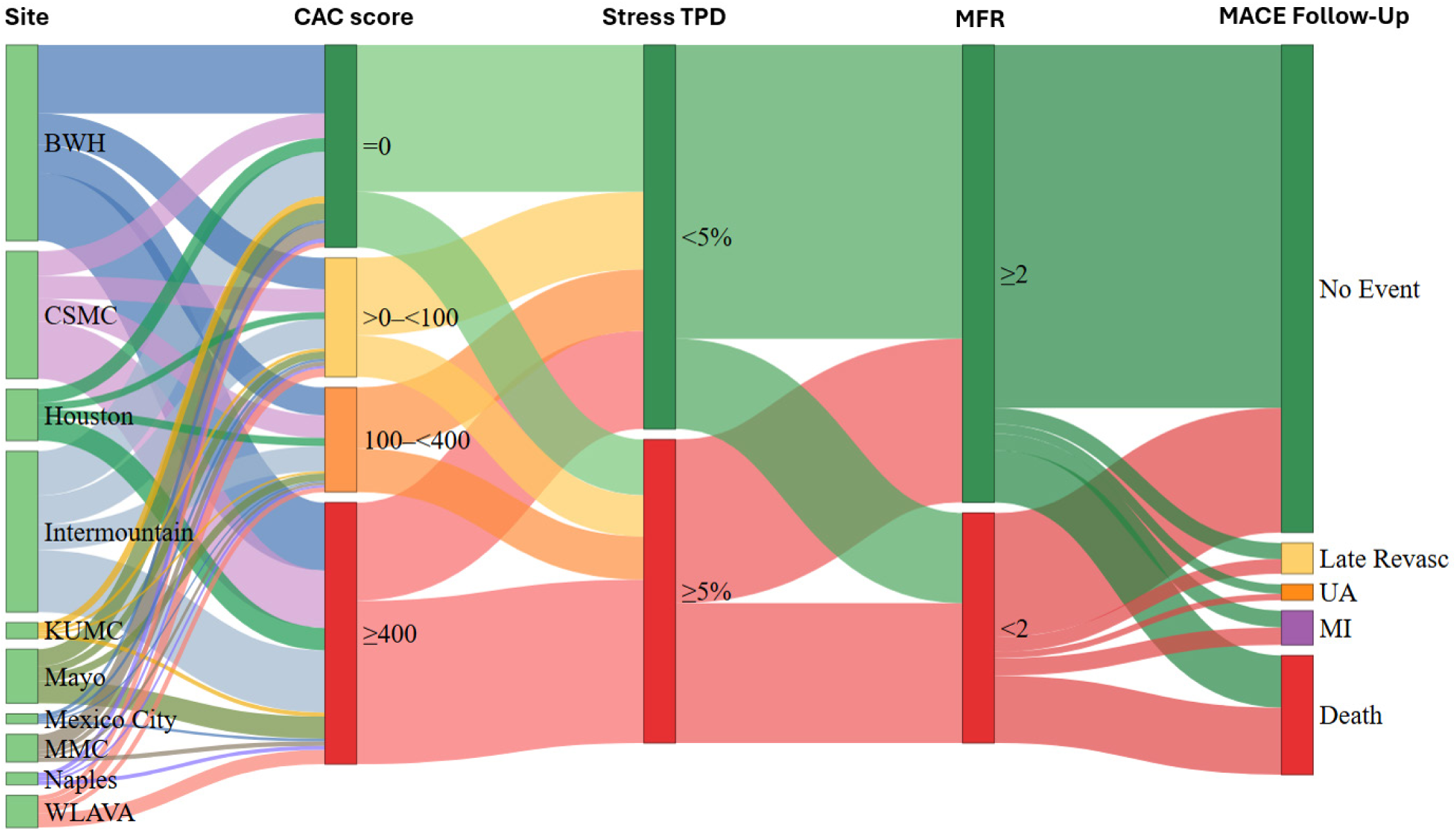
Sankey diagram illustrating data flow for the 10 sites with cleaned clinical and imaging data, including stress prefusion, dynamic imaging, and a CTAC scans. Rest CTAC scan was used when available; if stress-only imaging was performed, the stress CTAC scan was used. Total n= 25,310. CAC: coronary artery calcium; CTAC: computed tomography attenuation correction; TPD: total perfusion deficit; MFR: myocardial flow reserve; MACE: major adverse cardiovascular event; BWH: Brigham and Women’s Hospital; CSMC: Cedars-Sinai Medical Center, KUMC: University of Kansas Medical Center, Mexico City: National Autonomous University of Mexico, Naples: University of Naples Federico II, WLAVA: West Los Angeles Veterans Affair MCE

**Table 5** summarizes site-adjudicated perfusion and CAC data for patients with cleaned clinical data and imaging data that had been QC-ed by an expert in the core laboratory.

**Table 5:** Visual Perfusion Summary

| **Variable** | **Overall**<br>N = 22,212<sup>1</sup> | **Normal**<br>N = 12,780<sup>1</sup> | **Probably**<br>**Normal**<br>N = 1,230<sup>1</sup> | **Equivocal**<br>N = 896<sup>1</sup> | **Abnormal**<br>N = 2,443<sup>1</sup> | **Definitely**<br>**Abnormal**<br>N = 4,863<sup>1</sup> | **p-value<sup>2</sup>** |
| --- | --- | --- | --- | --- | --- | --- | --- |
| **Stress**<br>**TPD (%)** | 4.2 (1.6,<br>10.3) | 2.3 (0.9, 4.7) | 3.5 (1.9, 6.3) | 4.1 (2.7, 6.2) | 9.2 (5.0, 16.0) | 16.8 (9.5, 27.1) | <0.001 |
| **Ischemic**<br>**TPD (%)** | 3.0 (1.3, 6.2) | 1.8 (0.8, 3.5) | 2.6 (1.5, 4.3) | 3.2 (2.1, 4.6) | 5.7 (3.4, 9.1) | 8.7 (5.2, 14.2) | <0.001 |
| **Stress**<br>**EF (%)** | 67.3<br>(55.5, 75.2) | 71.5<br>(63.6, 77.9) | 68.6<br>(60.2, 74.8) | 67.9<br>(58.9, 74.4) | 61.8<br>(47.5, 70.4) | 52.7<br>(38.5, 64.2) | <0.001 |
| **Stress**<br>**MBF (%)** | 2.3 (1.7, 2.9) | 2.5 (2.0, 3.1) | 2.5 (1.9, 3.0) | 2.2 (1.8, 2.9) | 1.8 (1.4, 2.4) | 1.6 (1.2, 2.1) | <0.001 |
| **CAC**<br>**score** | 131<br>(0, 950) | 33<br>(0, 405) | 33<br>(0, 421) | 258<br>(21, 1035) | 518<br>(40, 1,773) | 826<br>(143, 2124) | <0.001 |
| **MFR** | 2.3 (1.8, 2.9) | 2.5 (2.1, 3.1) | 2.4 (1.9, 3.1) | 2.2 (1.7, 2.8) | 2.1 (1.6, 2.7) | 1.8 (1.4, 2.4) | <0.001 |

<sup>1</sup>Median (Q1, Q3)
<sup>2</sup>Kruskal-Wallis rank sum test

## DISCUSSION

We established a large international imaging registry of 35,588 patients, assembling a rich set of 110 clinical variables, 42 imaging variable categories, and over 2,200 individual imaging variables, along with complete DICOM data (encompassing CT, stress/rest static, gated, and dynamic PET) and clinical outcomes. All imaging variables were automatically quantified and merged with the clinical data, utilizing established clinical software and novel AI methods. These results include advanced metrics as subendocardial perfusion and flow. We integrated imaging data from multiple vendors with related clinical information. While this process can be automated, substantial differences between sites made it a labor-intensive task, requiring the collaboration of a dedicated team of scientists, clinicians, and research associates. This registry will enable validation, enhancement, and streamlining of both standard and novel cardiac PET/CT processing methods.

From a technical standpoint, organizing DICOM data presented a major challenge. While the lessons learned during the formation of the REFINE SPECT Registry helped us to avoid many hurdles, the high resolution of PET/CT MPI, with multiple acquisitions and reconstructions, made this task complex [10, 17]. Potential for missing, duplicative, or corrupted images –requires rigorous quality control measures to ensure accurate and reliable imaging outcomes. All cases were carefully QC-ed before the quantitative analysis. To avoid case mismatch, we routinely cross-linked the DICOM header data with clinical variables in the master spreadsheets. This approach enabled us to automatically identify discrepancies, such as mismatches in patient sex or age.

The diverse baseline clinical and imaging characteristics highlight the uniqueness of the registry for a more universal implementation of PET in clinical practice. The high prevalence of diabetes and obesity translates into a substantial number of patients who, despite a CAC score of 0, present with an MFR <2 [3, 6, 21]. Similarly, it was not uncommon to observe patients with advanced coronary atherosclerosis (CAC >400) but no perfusion or flow abnormalities. These patterns can be seen in **Figure 2**. Such discordance in the manifestation of CAD requires a comprehensive multimodality approach that can evaluate both the presence and extent of disease (anatomy), as well as the functional significance of coronary stenoses (perfusion and flow) – a capability that can be achieved with cardiac PET [22–24]. Indeed, to effectively guide medical and invasive therapies for CAD, it is essential to reliably and objectively quantify disease extent and severity. With its inherent multimodality capabilities, PET/CT MPI is uniquely positioned to address this clinical need. One area of development which can benefit from REFINE PET registry is streamlining management of image quality (9). For example, we aim to address the impact of patient motion, which can result in misregistration artifacts and reduced accuracy of MBF measurements. Furthermore, there is significant potential to enhance automation of both left and right ventricles [25, 26]. CTAC also, could serve as a valuable source of auxiliary data, which, if extracted automatically, could further streamline the process of image interpretation. Indeed, we have already developed and validated several novel applications of PET CTAC scans using a subset of the collected data [25, 27–31].

There are several limitations that should be considered. As with other observational registries, there are selection biases for patients referred for PET/CT MPI imaging. There is also potential heterogeneity between centers, in addition to inter-observer and inter-center variability in visual interpretation. However, this limitation was mitigated by the availability of image data with objective quantitative analysis. Coronary stenosis on ICA was assessed by visual assessment at each site, a method known to overestimate the severity of angiographic disease when compared with fractional flow reserve [32]. Fractional flow reserve measurements are not available in this population. The accuracy of stenosis interpretation may also differ between centers. The collected MACE events include all-cause death since the definition of cardiac death can be very difficult, particularly among elderly patients with multiple comorbidities.

## CONCLUSION

The REFINE PET registry provides a robust dataset integrating imaging, clinical and deep learning data. It will enhance our understanding of the clinical utility of PET, promote the use of multiparametric data contained within the integrated PET/CT MPI dataset, and advance the development and validation of artificial intelligence methods for enhancing diagnostic accuracy and risk stratification.

## Data Availability

To the extent allowed by data sharing agreements and IRB protocols, the deidentified data and data analysis code from this manuscript will be shared upon written request.

## ACKNOWLEDGEMENTS

We would like to thank all participating institutions and investigators who contributed clinical and imaging data to the REFINE PET registry. We are especially grateful to the data managers, research coordinators, and technical staff at each site for their critical role in the data collection. We acknowledge the support of the core laboratory at Cedars-Sinai Medical Center for data harmonization, imaging analysis, and infrastructure support. Special thanks to the imaging technologists, software engineers, and data scientists who developed and applied the automated artificial intelligence tools used in this study.

## DISCLOSURES

RM received consulting fees from Pfizer and research support from Pfizer and Alberta Innovates. DB and PS participated in software royalties for QPS software at Cedars-Sinai Medical Center. DB, DD, and PS reported equity in APQ Health Inc. DB received research grant support from The Dr. Miriam and Sheldon G. Adelson Medical Research Foundation and consulting fees from GE Healthcare. PS received research grant support from Siemens Medical Systems, and consulting fees from Synektik SA and Novo Nordisk. PC reported consulting for Clario. MDC reported consulting fees from MedTrace, Valo Health, GE, Bitterroot Bio, and IBA, investigator-initiated research support from Amgen, and institutional research grant support from Sun Pharma, Xylocor, Alnylam, and Intellia. AJE reported consulting for Artrya, authorship fees from Wolters Kluwer Healthcare—UpToDate and serving on scientific advisory boards for Axcellant and Canon Medical Systems USA; his institution has grants/grants pending from Alexion, Attralus, BridgeBio, Canon Medical Systems USA, GE HealthCare, Intellia Therapeutics, International Atomic Energy Agency, Ionis Pharmaceuticals, National Institutes of Health, Pfizer, and Shockware Medical. RRSP serves as a consultant for GE HealthCare. RS reported being a consultant for GE Healthcare. MA-M received research support from Siemens and GE Healthcare and is a consultant to Jubilant, Medtrace, GE Healthcare, and Pfizer. LS received grant support/consulting honorarium from Amgen and Philips and served as site PI for V-INITIATE and Ocean(a) trials. The remaining authors declare no conflict of interest.

### Independent Data Access and Analysis

GR, ML, AS, and PS had full access to all the data in the study and take responsibility for its integrity and the data analysis.

