## Supplementary Material for "The REgistry of Flow and Perfusion Imaging for Artificial INtelligEnce with PET (REFINE PET): Rationale and Design"

**SUPPLEMENTAL TABLES**

**Supplementary Table 1:** CT imaging parameters per site

| Site | Camera | Slice Thickness [mm] | Tube Current [mA] | Tube Voltage [kVp] |
|---|---|---|---|---|
| **Brigham and Women's Hospital** | GE Discovery MI<br>GE Discovery RX<br>GE Discovery STE | 2.5-5 | 10-26 | 120-140 |
| **Cedars-Sinai Medical Center** | Siemens Biograph 64 TruePoint<br>Siemens Biograph 128 Vision Edge<br>GE Discovery 710 | 3 | 11-13 | 100 |
| **Mayo Clinic** | GE Discovery 710 | 3.75 | 17-77 | 120 |
| **Intermountain Medical Center** | Siemens Biograph 16 TruePoint<br>Siemens Biograph 20 mCT<br>Siemens Biograph 40 mCT | 2 | 15-38 | 120-130 |
| **Montefiore Medical Center** | Philips Gemini TF TOF 16<br>Philips Gemini TF TOF 64 | 3 | 110-185 | 120 |
| **University of Kansas Medical Center** | GE Discovery MI | 3.75 | 14-75 | 120 |
| **University of Naples Federico II** | Philips Ingenuity TF<br>GE Discovery MI | 3 | 35-100 | 120-140 |
| **West Los Angeles Veterans Affairs Medical Center** | Siemens Biograph 64 mCT<br>Siemens Biograph 64 Vision 600 | 3 | 70-200 | 120 |

| | | | | |
|---|---|---|---|---|
| **Columbia University Irving Medical Center** | Siemens Biograph 64 mCT Flow | 3 | 75 | 120 |
| **Houston Methodist Academic Institute** | Siemens Biograph 600 Vision Edge | 3 | 20-50 | 100-120 |
| **Ottawa Heart Institute** | GE Discovery 690<br>GE Discovery 600<br>Siemens Biograph 600 Vision Edge | 3-5 | 20-60 | 120 |
| **Rush University Medical Center** | Siemens Biograph Horizon | 5 | 40-120 | 110 |
| **National Autonomous University of Mexico** | Siemens Biograph 64 TruePoint<br>Siemens Biograph Vision 600 | 3 | 33-580 | 120 |
| **University Hospital Zurich** | GE Discovery STE<br>GE Discovery RX<br>GE Discovery LS<br>GE Discovery HR | 3.75-5 | 140 | 40-240 |
